# Prevalence and demographic distribution associated with pre-eclampsia among pregnant women at a local Teaching Hospital in Ghana

**DOI:** 10.1101/2022.05.18.22275250

**Authors:** Nyarko Adwoa, Joshua A. Kunfah, Collins Adombine Akayuure, Jamilatu Kappiah, Sylvanus Kampo

## Abstract

**Background:** In sub-Saharan Africa, pre-eclampsia remains a major health problem contributing to high rates of maternal mortality. Despite this condition having adverse effects on maternal and child health, its prevalence and associated risk factors are still significant, especially in developing countries including Ghana. This study aimed to assess the prevalence and demographic distributions associated with pre-eclampsia among pregnant women at the Ho Teaching Hospital.

**Methods:** A facility-based retrospective study was conducted by reviewing available data or hospital records of pregnant mothers who were admitted to the labour and maternity wards from January 2018 to December 2020. The data were collected using a structured checklist.

**Results:** A total of 5,609 deliveries were recorded from 2018 to 2020. Out of the 5,609 deliveries, 314 pre-eclampsia cases were recorded giving an overall prevalence of 5.6 %. The yearly prevalence for 2018, 2019, and 2020 were 4.6 %, 5.6 %, and 6.6 %, respectively. The most recorded pre-eclampsia cases were seen among women within the age group of 18-24 years. The data showed that 112 (35.7 %) of the mothers who had pre-eclampsia were nulliparous. Pre-eclampsia associated maternal and fetal complications were; preterm delivery 221 (70.4 %), intrauterine fetal death 62 (19.7 %), eclampsia 9 (2.9 %), HELLP syndrome 5 (1.6 %) and maternal death 17 (5.4 %). Associated factors of pre-eclampsia were parity, level of education, and occupation (p=<0.05).

**Conclusion:** The findings of this study showed a rising trend in the incidence of pre-eclampsia over the years at the Ho Teaching Hospital. Parity, level of education, and occupation were found to be associated with developing pre-eclampsia.

## Introduction

Pre-eclampsia remains a major clinical challenge in contemporary obstetric practice due to its associated burden of high maternal and perinatal adversities (Adu-Bonsaffoh et al., 2017). It is multi-factorial and forms an integral part of the continuum of hypertensive disorders in pregnancy [HDP] (Kooffreh et al., 2014). They are common medical conditions in pregnancy responsible for approximately 14 % of maternal deaths globally (Jane R, Hans M, Ali S, 2020).

In Ghana, the maternal mortality ratio remains excessively high and hypertensive disorders in pregnancy is responsible for about 9 % of maternal deaths. However, recent clinical studies in Ghana have indicated that HDP is the leading cause of maternal deaths in the major tertiary institutions with similar findings reported in other African countries (Adu-Bonsaffoh et al., 2017). Pre-eclampsia is a fore-arm of eclampsia, a pregnancy-specific syndrome characterized by new onset of hypertension and significant protein in urine with or without edema occurring at 20 weeks of gestation. It is associated with high maternal mortality and morbidity as well as the risk of fetal perinatal death, preterm birth, intrauterine growth restriction, placenta abruption, oligohydramnios and other pathology (Otu-Nyarko et al., 2015). However, it is mostly asymptomatic and difficult to predict in the early stages of pregnancy. As a result, most cases are not detected early and seen at health facilities leading to severe eclampsia (high blood pressure in pregnancy with or without protein in urine). Pre-eclampsia is the second leading cause of maternal death and it has been associated with maternal morbidity and adverse perinatal outcome globally though there is no known treatment except the delivery of the placenta (Frank et al., 2020; Otu-Nyarko et al., 2015).

According to World Health Organization (WHO), its incidence is seven times higher in developing countries(2.8% of live births) than in developed countries [0.4%] (Dolea & Abouzahr, 2003). In the United States of America, pre-eclampsia is believed to be responsible for 15% of premature deliveries and 17.6% of maternal deaths (Creanga et al., 2017). Despite its impact on maternal and child health, efforts to predict and prevent pre-eclampsia have been disappointing and remain one of the poorly understood obstetric complications with its adverse effect on maternal and child health (Moussa et al., 2014). Its prevalence is still significant, especially in developing countries including Ghana and yet a major threat to maternal and neonatal health. Hence, this study aimed to access the prevalence, demography distribution and associated risk factors of pre-eclampsia among pregnant women living in the Ho municipality.

## Methods

A hospital-based retrospective study with a quantitative approach was utilized to assess the prevalence and social demographic factors associated with pre-eclampsia using past medical information of pregnant women at the Ho teaching hospital.

The study was conducted over a period of two months. The ethical committee of the University of Health and Allied Sciences approved the study protocol (UHAS-REC A.12[177]20-21). The study retrieved all pregnant women within the age group of eighteen (18) and above who delivered in the facility from January 2018 to December 2020 using the Hospital Administration and Management System (HAMS). All pregnant women who were diagnosed with pre-eclampsia within this period were included in the data. Data was collected using a checklist which assessed the socio-demographic variables including; patient age, marital status, weight, height, residence, occupation, religion, educational level, and parity. The maternal age of the patient was classified into three categories (18-25, 25-35and ≥35 years), marital status was dichotomized into married and unmarried, maternal educational level grouped into primary, secondary and tertiary levels, and maternal body mass index (BMI) as maternal weight in kilograms divided by maternal height in meters square and obstetric related factors grouped as gravidity, parity, gestational age.

### Statistical Analysis

The complete data in Microsoft Excel was exported to Statistical Package for Social Science (SPSS) version 22.01 (IBM Corporation, Armonk, NY, USA) which was used for data entry and analysis. Mean and SD was computed for quantitative variables such as age, weight, gestational age, and BMI. Descriptive analysis was done to evaluate the distribution of variables and statistical findings were reported as numbers, percentages, and frequencies. Binary analysis was done to determine the potential demographic associated with pre-eclampsia. The association between independent and dependent variables was measured and tested using chi-square and a P-value of ≤ 0.05 was considered significant.

## Results

### Socio-demographic Profile of Pre-eclampsia Cases

A total of 5,609 deliveries were recorded at the Ho Teaching Hospital from January 2018 to December 2020. The mean age of the respondents was 28.89. The data showed that 314 respondents out of the 5,609 deliveries were diagnosed with pre-eclampsia, 27.7% of the pre-eclampsia cases were within the age group of 18-24 years old, 25.5% of them were within 25-29 years old, 26.1 % were within the age group of 30-34 years old, whereas 4% of them were over 40 years old. The data also showed that 93 % of the pre-eclampsia cases were Christian, whereas 7% of them were Muslims. The results showed that 67.5% of the pre-eclampsia cares were married and 32.5 % of them were not married. Regarding their level of education, the data showed that 38.9 % of the pre-eclampsia patients had Junior High School education, whereas 4.5% of them had no education **(Table 1)**.

**Table-1:** Socio-demographic Characteristics of pre-eclampsia cases.

| Variable | Frequency | Percentages (%) |
| --- | --- | --- |
| **Age** |  |  |
| 18-24 | 87 | 27.7 |
| 25-29 | 80 | 25.5 |
| 30-34 | 82 | 26.1 |
| 35-39 | 48 | 15.3 |
| 40-44 | 15 | 4.8 |
| 45-49 | 2 | 0.6 |
| **Marital status** |  |  |
| Married | 213 | 67.8 |
| Single | 101 | 32.2 |
| **BMI** |  |  |
| <18.5 | 11 | 3.5 |
| 18.5-24.9 | 143 | 45.6 |
| 25.0-29.9 | 153 | 48.7 |
| 30.0-35.0 | 7 | 2.2 |
| **Level of education** |  |  |
| No Education | 14 | 4.5 |
| Primary | 73 | 23.2 |
| JHS | 122 | 38.9 |
| Secondary | 67 | 21.3 |
| Tertiary | 38 | 12.1 |
| **Religion** |  |  |
| Christians | 292 | 93.0 |
| Muslims | 22 | 7.0 |
| **Occupation** |  |  |
| Farming | 32 | 10.2 |
| Government Workers | 51 | 16.2 |
| Student | 47 | 15 |
| Trading | 155 | 49.4 |
| Unemployed | 29 | 9.2 |

We investigated the prevalence rate of pre-eclampsia in Ho Municipality from 2018 to 2020. The data showed a prevalence rate of 5.6 % for 5,609 deliveries. We also determined the yearly prevalence rate of pre-eclampsia and observed a rate of 4.6 % in 2018 for 1950 deliveries, 5.6 % in 2019 for 1,809 deliveries, and 6.6% in 2020 for 1,850 deliveries **(Table 2)**.

**Table 2:** Prevalence of pre-eclampsia from 2018 to 2020.

| **Year** | **Deliveries** | **preeclampsia cases** | **Percentage (%)** |
| --- | --- | --- | --- |
| 2018 | 1950 | 89 | 4.6 |
| 2019 | 1809 | 102 | 5.6 |
| 2020 | 1850 | 123 | 6.6 |
| **Total** | **5609** | **314** | **5.6** |

The occurrence of complications associated with pre-eclampsia from the study recorded that 70.4 % of the pre-eclampsia patients suffered from preterm delivery, 19.7 % of them had intrauterine fetal death, 2.9 % of them developed eclampsia, 1.6 % of them developed HELLP syndrome and 5.4 % of them resulted in maternal deaths **(Table 3)**.

**Table 3:** Distribution of pre-eclampsia complications among 314 pregnant women.

| **Complications**<br>**Of pre-eclampsia** | **Number of pregnant**<br>**women affected** | **Percentage (%)** |
| --- | --- | --- |
| Preterm delivery | 221 | 70.4 |
| Intra-uterine fetal<br>death(IUFD) | 62 | 19.7 |
| HELLP syndrome | 5 | 1.6 |
| Maternal death | 17 | 5.4 |
| Eclampsia | 9 | 2.9 |

We examined the distribution of pre-eclampsia cases within the Volta region for the period of 2018 to 2020 the cases recorded at the Ho Teaching Hospital. The data showed that 47 (15 %) of the pre-eclampsia cases were from Kpetoe, 26 (8.3 %) were from Peki, 24 (7.6) were from Hohoe, 22 (7 %) were from Adaklu and 19 (6.1 %) were from Adidome whilst Ho recorded the highest of 37.3% **(Table 4)**.

**Table 4:** Distribution of pre-eclampsia cases within the Volta Region from 2018 to 2020.

| Towns | Total deliveries | Pre-eclampsia cases | Percentage (%) |
| --- | --- | --- | --- |
| Adaklu | 393 | 22 | 7 |
| Adidome | 339 | 19 | 6.1 |
| Dzemeni | 286 | 16 | 5.1 |
| Dzodze | 322 | 18 | 5.7 |
| Ho | 2090 | 117 | 37.3 |
| Hohoe | 429 | 24 | 7.6 |
| Keta | 232 | 13 | 4.1 |
| Kpetoe | 840 | 47 | 15 |
| Peki | 464 | 26 | 8.3 |
| Sogakope | 214 | 12 | 3.8 |

The binary analysis from Tables 4 and 5 indicates the association between demographic factors and pre-eclampsia among pregnant women who were diagnosed with preeclampsia at Ho Teaching Hospital within the said period. There was no significant association between age, BMI, marital status, religion, address, and gravidity (P-value = > 0.05). However, there was a significant association between parity, level of education and occupation with a P-value of 0.004, 0.00 and 0.003 respectively **(Tables 5 and 6)**.

**Table 5:** Associations between Pre-eclampsia and demographic distributions.

| Variable | Total | Yearly Distributions |  |  | Chi-Square | p-value |
| --- | --- | --- | --- | --- | --- | --- |
|  |  | 2018 | 2019 | 2020 |  |  |
| **Age** |  |  |  |  | 10.306a | 0.414 |
| 18-24 | 87 | 33 | 27 | 27 |  |  |
| 25-29 | 80 | 23 | 26 | 31 |  |  |
| 30-34 | 82 | 17 | 29 | 36 |  |  |
| 35-39 | 48 | 12 | 15 | 21 |  |  |
| 40-44 | 15 | 4 | 5 | 6 |  |  |
| 45-49 | 2 | 0 | 0 | 2 |  |  |
| **BMI** |  |  |  |  | 1.699a | 0.945 |
| <18.5 | 11 | 3 | 3 | 5 |  |  |
| 18.5-24.9 | 143 | 41 | 49 | 53 |  |  |
| 25.0-29.9 | 153 | 44 | 48 | 61 |  |  |
| >=30 | 7 | 1 | 2 | 4 |  |  |
| **Marital status** |  |  |  |  | 5.362a | 0.068 |
| Married | 213 | 52 | 71 | 90 |  |  |
| Single | 101 | 37 | 31 | 33 |  |  |
| **Religion** |  |  |  |  | 1.502a | 0.472 |
| Christian | 292 | 81 | 94 | 117 |  |  |
| Muslim | 22 | 8 | 8 | 6 |  |  |
| **Level of education** |  |  |  |  | 47.473a | 0.000 |
| JHS | 122 | 18 | 39 | 65 |  |  |
| No Education | 14 | 7 | 7 | 0 |  |  |
| Primary | 73 | 37 | 21 | 15 |  |  |
| Secondary | 67 | 21 | 23 | 23 |  |  |
| Tertiary | 38 | 6 | 12 | 20 |  |  |
| **Occupation** |  |  |  |  | 23.446a | 0.003 |
| Farming | 32 | 13 | 8 | 11 |  |  |
| Government Workers | 51 | 6 | 16 | 29 |  |  |
| Student | 47 | 22 | 16 | 9 |  |  |
| Trading | 155 | 42 | 53 | 60 |  |  |
| Unemployed | 29 | 6 | 9 | 14 |  |  |

**Table 6:**
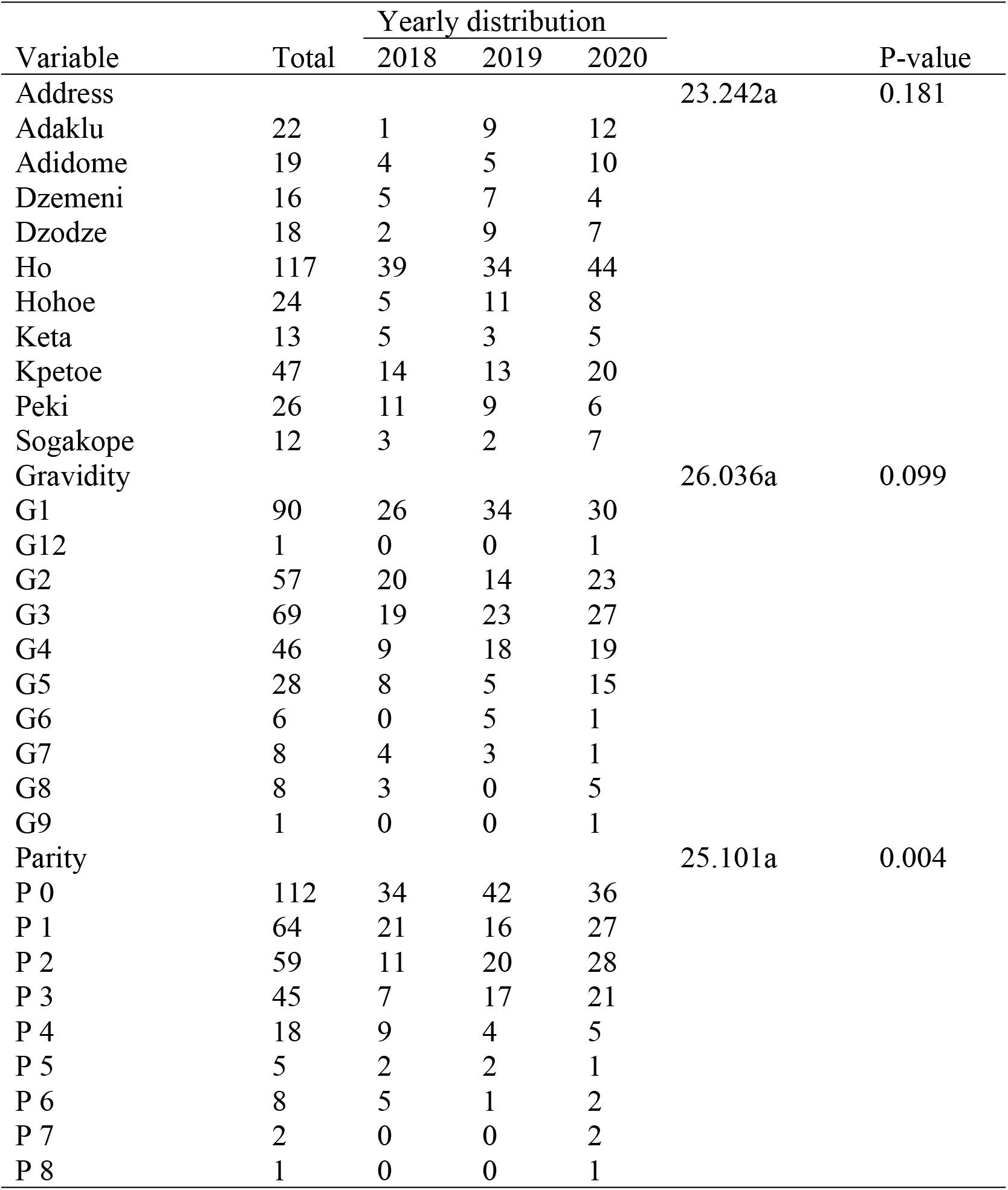
Association between Pre-eclampsia and demographic distribution factors.

## Discussion

### Prevalence of pre-eclampsia

The prevalence of pre-eclampsia over the period of study was 5.6 % which is higher compared to other studies reported in Norway (Klungsøyr et al., 2012), and Germany (Schneider et al., 2011) with 3 %, and 2.3 %, respectively. Our study fell below the estimated prevalence rate of 6.55 – 7.03% pre-eclampsia in a study conducted in Ghana (Ahenkorah, 2009). A further study conducted at the Korle - Bu Teaching Hospital (Obed & Patience, 2006) supports Ahenkorah estimated range with a prevalence rate of 7.03% in pre-eclampsia. This is a clear indication of how medical performance improved during the years as practitioners showed interest in understanding and reducing the complications of pregnancy, especially pre-eclampsia. The high rate of prevalence at the Korle-Bu Teaching Hospital could also be due to the high patient attendance since it is one of the largest referral centres in Ghana. However, this prevalence was also lower compared to studies conducted in Ethiopia and Nigeria with a prevalence rate of 12.4 % and 16 % respectively (Belay & Wudad, 2019b; Guerrier et al., 2013). Over the study period, there was a slight increase in the prevalence from; 4.6 % in 2018 to 5.6 % in 2019 and 6.6 % in 2020. The factors responsible for this slight increase are not clear in this study.

The difference between our findings and that of previous studies could also be due to the variations in socio-demographic characteristics of pregnant women. Fondjo et al (2019), stipulated that advanced age and age below 18 years of pregnant women are significantly predisposed to developing pre-eclampsia. Thus, the low prevalence rate observed in this study can be attributed to the fact that ages below 18 years were excluded from the sample and the highest rate of attendants were within the age group of 18 to 24 as well as few women above 45 years.

### Complications of Pre-eclampsia

Pregnant women who had pre-eclampsia in this study were identified with complications including; preterm delivery, intrauterine fetal death (IUFD), eclampsia, and HELLP syndrome. When poorly managed pre-eclampsia could progress into eclampsia which is characterized by seizures, in addition to exhibiting the symptoms of pre-eclampsia (Logan et al., 2021). The low recordings of pregnant women diagnosed with eclampsia could be the fact that medical personnel at the Ho Teaching Hospital put maximum effort into caring for pregnant women diagnosed with pre-eclampsia to prevent the progress of the condition into complications. Consistent with the findings of this study, Duley (2009), reported that up to about 20 % of preterm deliveries are a result of pre-eclampsia and have been documented to result in high neonatal mortalities and prolonged neonatal morbidities. According to Jeyabalan (2013), pre-eclampsia does not only result in neonatal mortalities, but accounts for up to about 9 % of maternal deaths in sub-Saharan Africa, and Asia. Consequently, adequate management of pre-eclampsia is the best means of preventing complications. It is noteworthy that although there have been several advances in medical science, the only known cure for pre-eclampsia is the delivery of the fetus and the placenta (Jeyabalan, 2013).

### Demographic Distribution and Association with Pre-eclampsia

The study identified a statistically significant association between parity and severity of Pre-eclampsia (P = 0.04) which is inconsistent with findings from a study conducted in Kenya, that nulliparous and primiparous women were at an increased risk of developing pre-eclampsia, compared to multiparous women (Logan et al., 2021). Grum et al. (2017), also reported an association between parity and the development of pre-eclampsia among pregnant women. Also, level of education had an association with pre-eclampsia (p=0.00). A study conducted by Fondjo et al., 2019 reported that pregnant women who had poor knowledge of pre-eclampsia stood at risk of being diagnosed with pre-eclampsia with the progress of their pregnancy.

In our study, women who completed their education at the junior high school level indicated an increased rise in pre-eclampsia than those who attained a higher educational level. The increased risk of pre-eclampsia among women with low education levels could be because, in Ho, people with low income are more likely to practice poor lifestyle practices including lack of physical exercise and poor eating habits that could lead to overweight or obesity which increases the risk of developing pre-eclampsia. On the other hand, the increased risk of pre-eclampsia among women with low education could be attributed to low maternal age, as women with reduced or less education achievement are more likely to have their children at an early maternal age. Similarly, a study conducted in Uganda has reported higher education to be protective against pre-eclampsia (Kiondo et al., 2012). This study also found occupations associated with pre-eclampsia (P = 0.003) with the majority of the pregnant women being traders. Jeyabalan (2013), in his study, indicated that increasing BMI is associated with an increased risk of developing pre-eclampsia in pregnancy which conflicts with our study findings. The differences could be due to the study designs that were utilized. A retrospective cross-sectional study was used in our research as Jeyabalan, 2013 utilized the case-control method which offered the opportunity to identify risk factors.

### Conclusion

The prevalence of pre-eclampsia was found to be rather low in this study, compared to rates that have been documented by some previous Ghanaian studies, and in other parts of Africa. But the study showed a rising trend in the incidence of pre-eclampsia over the years. Parity, level of education, and occupation were found to be associated with developing pre-eclampsia, whereas the remaining demographic characteristics showed no associations.

### Study Limitations

The limitation of the study was the small sample size analyzed due to problems with the retrieval of patients’ records and information. Also, the study design used in this study was a retrospective cross-sectional study, limiting the chances of identifying risk factors associated with pre-eclampsia. This study was conducted in a tertiary healthcare facility and the findings cannot be generalized in other healthcare settings. Further, because hospital records were used in collating data of pregnant women, some pertinent information could not be identified. In addition, pregnant women below 18 years of age were not included in this study, although past studies have reported pre-eclampsia among girls below 18 years of age.

### Recommendation

Further studies should be conducted to identify the risk factors associated with pre-eclampsia among this population. This is important to channel educational interventions in the right direction. In addition, the diagnosis of pre-eclampsia was determined by written reports in hospital records. It is unclear the means of diagnosis of pre-eclampsia, offering the possibilities of missed diagnosis and misdiagnosis. Consequent studies should endeavour to evaluate all markers for diagnosis for onward inclusion into the sample. Again, because of the challenge of paucity of data, hospital personnel should be encouraged to collate detailed information about patients before their being attended to at the facility. This would allow for more objective policies to be formulated, and directed towards achieving specific goals. In addition, it is pertinent for all females of reproductive age to be included in future studies, as opposed to this study where only females above 18 years of age were utilized.

## Data Availability

All data relevant are included in the manuscript and supporting documents.

## DECLARATIONS

### Ethics approval and consent to participate

The ethical committee of the University of Health and Allied Sciences approved the study protocol (UHAS-REC A.12 [177]20-21). Written informed consent was obtained from each recruited parturient after providing them with adequate explanations regarding the aims of this study.

### Consent to publish

Not applicable

### Availability of data and materials

The datasets generated and/or analyzed during the current study are not publicly available due to patient confidentiality but are available from the corresponding author on reasonable request.

## Authors’ contributions

NA and SK conceived and designed the study. SK was responsible for the supervision and coordination of this study. NA, JAK, CAA, JK and SK conducted the data collection. NA and SK led the data analysis with inputs from JAK, JAK, and CAA. JAK wrote the first draft of the manuscript, and then NA, JAK, CAA, JK and SK contributed to revising and reviewing the manuscript. All authors read and approved the final manuscript before submission.

## Acknowledgements

We thank the Chief Executive Officer (CEO) of the Ho Teaching Hospital and the staff of the department of obstetrics and Gynecology, at Ho Teaching Hospital for making available all the necessary materials needed for this study.

